# Lifetime Risk of Injury-Related Hospitalization in Canada: A Rough Estimate

**DOI:** 10.64898/2025.12.26.25343067

**Authors:** Qizhi Mao, Brandy Tanenbaum, Shaelyn Fitzpatrick

## Abstract

**Background:** Injury is one of the leading causes of hospitalization among Canadians and represents a substantial public health burden. However, at the population level, the lifetime risk of injury-related hospitalization is unknown. Estimating the lifetime risk of hospitalization due to injury among Canadians provides an intuitive and policy-relevant metric to inform the development of injury prevention strategies and health system planning.

**Methods:** This study used the Cumulative Risk Method to estimate the lifetime risk of at least one injury-related hospitalization among Canadians. Age-specific risks were calculated and summed under assumptions of a stable population and constant injury incidence. Estimates were adjusted for an average of 1.4 hospitalizations per person, and sensitivity analyses assessed the impact of varying this assumption.

**Results:** The estimated lifetime risk of experiencing at least one injury-related hospitalization among Canadians was 42.0% (95% CI: 41.9% - 42.1%), corresponding to approximately one in every 2.4 individuals over the life course. Lifetime risk was slightly higher among females (42.1%, 95% CI: 42.0% - 42.3%) than males (41.3%, 95% CI: 41.1% - 41.4%), although males exhibited higher cumulative risk before age 65. Sensitivity analyses indicated that, under different assumptions regarding repeat hospitalizations, lifetime risk estimates ranged from 31.1% to 53.5%.

**Discussion:** A substantial proportion of Canadians will experience at least one injury-related hospitalization over their lifetime. Estimates of lifetime risk provide a clear and easily interpretable metric that can communicate the burden of injury to the general population while highlighting to healthcare providers the importance of implementing sustained, population-level injury prevention strategies.

**What is already known on this topic:** Injury is a leading cause of hospitalization and death among Canadians, especially those under 45, and imposes a substantial healthcare and economic burden.

**What this study adds:** This study estimates that one in every 2.4 Canadians will experience one or more injury-related hospitalizations over their lifetime. It also provides sex-specific differences and examines the impact of repeated hospitalizations on risk.

**How might this study affect research, practice, or policy:** These findings provide a clear and easily understood measure of injury burden, which can guide public health planning, inform prevention strategies, and raise public awareness of the lifetime risk of serious injury.

## Background

Injury is the leading cause of death for Canadians under the age of 45 years, with an estimated cost of $29.4B annually^1^. Despite the health and financial toll of preventable injury, investment in prevention strategies and resources remains low. Injury-related hospitalizations impose a heavy burden on both individuals and society, higher than that from any other cause^2^. In 2018– 2019, a total of 225,208 people in Canada were hospitalized due to injuries; injury-related hospitalizations ranked second among children aged 1–9 and eighth across all age groups^1^. The financial impact is substantial, with more than $6.4B attributed to injury-related hospitalizations^2^. For healthcare providers, it is important to understand the risk of injury-related hospitalization and to adopt cost-effective intervention strategies.

Public health specialists commonly use indicators such as incidence and prevalence to describe the risk and burden of a disease. Lifetime risk is frequently expressed as a percentage or as ‘1 in X individuals,’ a format that is more easily understood and noticed by the general population. In the Netherlands, it is estimated that 1 in every 2.3 male infants will develop cancer at some point during their life; globally, this figure is approximately one in four^3,4^. In England, approximately 60% of people will undergo surgery in their lifetime^5^. In Canada, while some studies have assessed the lifetime risk of hip fractures, there is a lack of research estimating the lifetime risk due to injuries from all causes^6^.

Injury-related hospitalization is selected as the metric in this study to distinguish between lower acuity injuries that may require no medical intervention, a brief primary physician appointment or emergency department visit, and those injuries that meet a threshold of severity requiring hospital admission. These may range from single-body system injury to multi-body system trauma, and may include temporary or permanent, partial or full disability. Hospitalization is a standard measurement across regions and time, representing higher personal and system costs, making it a clear and actionable indicator for prevention efforts.

This study aims to estimate the lifetime risk of injury-related hospitalization among Canadian infants using the Cumulative Risk method, which has been previously validated and described by Schouten and his colleagues^7^.

## Method

### Data source

Data from the 2021 Census of Population were used to calculate the total population by age group and sex^8^. The number of hospitalizations due to injury by age group and sex for 2021– 2022 was obtained from the CIHI database^9^. Owing to the unavailability of detailed age stratification within the 18–64-year cohort in the CIHI database, this study incorporates injury-related hospitalization risk data by age group for individuals aged 18 to 64 derived from the 2018–2019 period as reference^10^.

### Statistic analysis

We performed data processing and visualization using R version 4.4.1. Using the Cumulative Risk Method, we roughly estimated the lifetime risk of injury-related hospitalization among Canadians, along with 95% confidence intervals (CIs)^7^. We estimated age-specific risks by dividing the number of injury hospitalizations in each age group by the corresponding population from the 2021 census. We assumed that population size, population structure, and age-specific injury risks would remain stable.

### Derivation of variables

To eliminate the confounding effect of readmissions on the calculations, we introduced the parameter *m*, representing the average number of hospitalizations due to injury. This parameter is difficult to estimate directly, but some previous studies have shown that among elderly patients hospitalized due to falls, 44.6% are readmitted within one year^11^; nearly one-third of patients injured in motor vehicle collisions experience a subsequent motor vehicle collision in America^12^; and the overall one-year readmission rate for trauma patients is 21.1% in Canada^13^. Considering that most studies only assess the one-year readmission rate, the actual lifetime readmission rate is likely to be higher. Under the conservative assumption of a 40% readmission rate, the parameter *m* is therefore 1.4. Assuming that an 85+ person has an average of 3 more years to live, and the total population size, population structure, and incidence rate remain stable over the subsequent years.

### Sensitivity analysis

We conducted a sensitivity analysis by varying the average number of hospitalizations due to injury (m) and repeated the analyses. This was done to examine the impact of readmission frequency on the estimation of lifetime risk.

## Results

Based on data from the 2021 Canadian Census, the total population was 36,991,980, comprising 18,765,735 females and 18,223,245 males. In the 2021–2022 period, a total of 272,761 individuals were hospitalized due to injury, including 144,078 females and 128,683 males. The distribution of injury-related hospitalizations across different age groups is presented in Table 1.

**Table 1:**
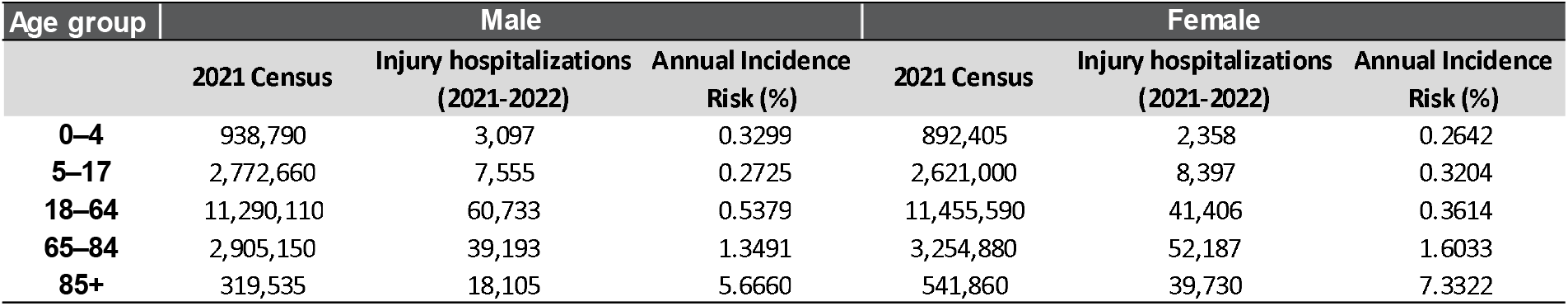
Annual incidence risk (%) of injury-related hospitalization by age group and sex.

| Age group | Male |  |  | Female |  |  |
| --- | --- | --- | --- | --- | --- | --- |
|  | 2021 Census | Injury hospitalizations (2021-2022) | Annual Incidence Risk (%) | 2021 Census | Injury hospitalizations (2021-2022) | Annual Incidence Risk (%) |
| 0–4 | 938,790 | 3,097 | 0.3299 | 892,405 | 2,358 | 0.2642 |
| 5–17 | 2,772,660 | 7,555 | 0.2725 | 2,621,000 | 8,397 | 0.3204 |
| 18–64 | 11,290,110 | 60,733 | 0.5379 | 11,455,590 | 41,406 | 0.3614 |
| 65–84 | 2,905,150 | 39,193 | 1.3491 | 3,254,880 | 52,187 | 1.6033 |
| 85+ | 319,535 | 18,105 | 5.6660 | 541,860 | 39,730 | 7.3322 |

Assuming (m = 1.4), the estimated lifetime risk of injury-related hospitalization among Canadian newborns is approximately 42% (95% CI: 41.87%–42.12 %), indicating that, on average, one in every 2.38 newborns is expected to experience injury-related hospitalization at some point during their lifetime. When stratified by sex, the lifetime risk is 41.25% (95% CI: 41.07%– 41.43%) for males, slightly lower than 42.11% (95% CI: 41.95%–42.28%) for females, with the difference being statistically significant. Notably, before the age of 65, males have a higher lifetime risk of injury-related hospitalization than females, with the greatest disparity observed during adulthood, indicating that cumulative risk among males substantially exceeds that of females from adulthood through to older age.

In the sensitivity analysis for parameter *m*, we considered two extreme scenarios. First, assuming that each individual is hospitalized for injury only once in their lifetime, the estimated lifetime risk of hospitalization is 52.5% for men and 53.5% for women. Second, if all individuals who have experienced an injury-related hospitalization eventually experience a second hospitalization, the estimated lifetime risk of hospitalization is 31.1% for men and 31.8% for women. These results are summarized in Figure 2.

**Figure 1.**
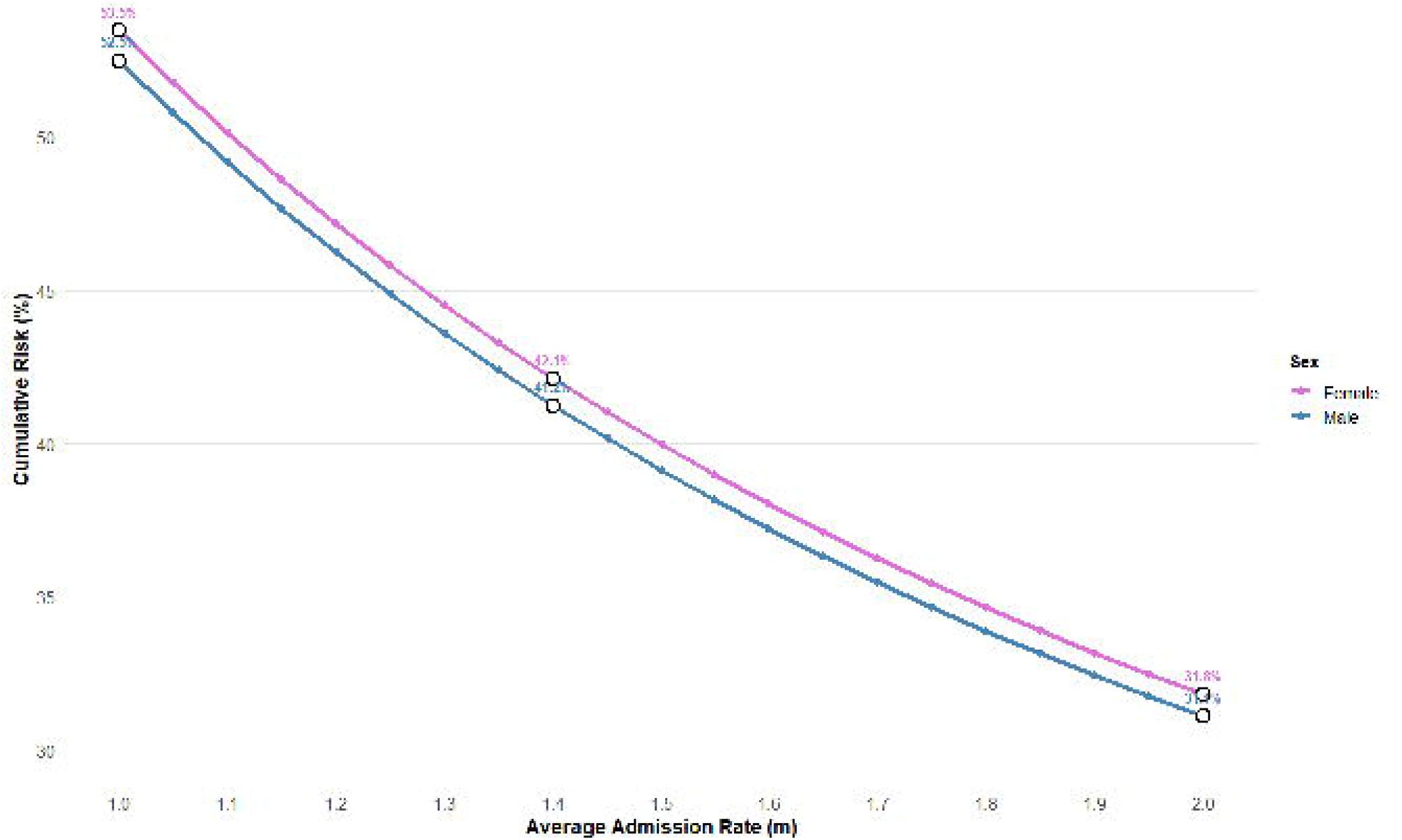
Age- and sex-specific distribution of injury-related hospitalizations and cumulative risk across the lifespan in Canada (m=1.4).

**Figure 2.**
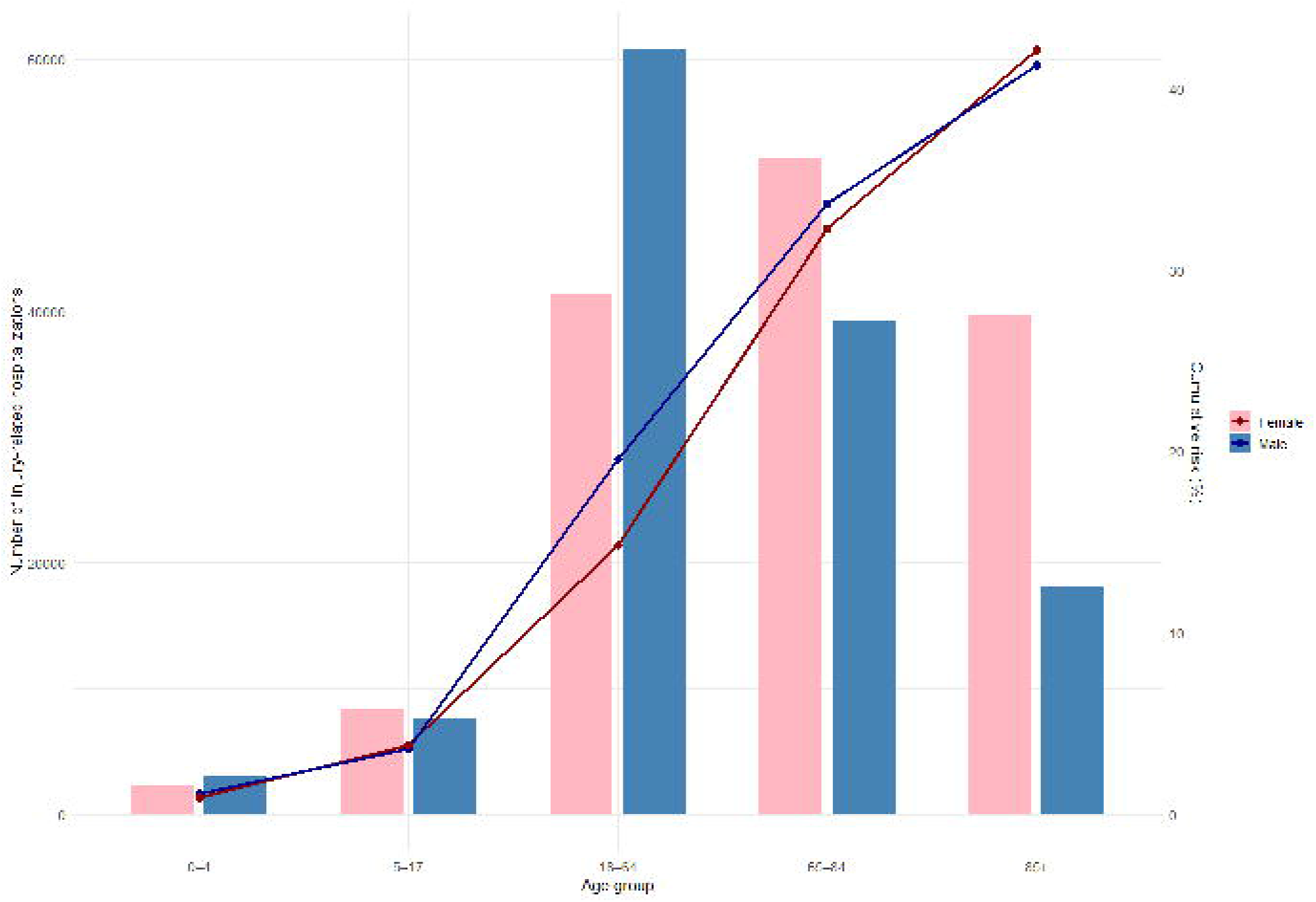
Sensitivity analysis stratified by sex.

## Discussion

This study provides the first estimate of the lifetime risk of injury-related hospitalization among Canadian newborns, suggesting that approximately 42%, or one in every 2.4, may experience at least one hospitalization due to injury over their lifetime. These findings highlight that injuries are not isolated events but represent a substantial public health concern with significant implications for both individuals and the healthcare system. Unlike previous research that focused on specific injury types, this study used all-cause injury hospitalization data, allowing for a more comprehensive assessment of the overall burden.

Using the Cumulative Risk Method combined with the most recent census and hospitalization data, we derived an intuitive metric for policymakers and the public. Sex-stratified analyses indicated a marginally higher lifetime risk among females compared to males (difference ≈0.9%), suggesting that sex-specific considerations may be relevant when designing prevention strategies, although the overall difference is minimal, emphasizing the importance of broad-based preventive measures.

Importantly, injury risk is not experienced equally across a diverse population. Previous research suggests that those most vulnerable, with higher marginalization and social needs, experience a greater risk of injury and reinjury^14^. Beyond age and sex, this study does not account for social or biological factors that influence health outcomes, including injury. When health equity data becomes available, an updated analysis could help to understand risk using an intersectional approach to identify differences within the population and strategize resources and interventions accordingly.

Sensitivity analyses confirmed the robustness of the estimates with respect to the parameter *m* (average number of hospitalizations), indicating that the results provide a reasonable approximation of lifetime risk despite inherent uncertainties. Nevertheless, several limitations warrant consideration. The Cumulative Risk Method may overestimate risk, and the absence of detailed age-stratified data for adults precluded the use of life-table approaches. Additionally, the analysis did not distinguish between specific injury mechanisms (e.g., falls, motor vehicle collisions) or severity, potentially masking important heterogeneity in risk.

Despite these limitations, the findings offer a quantitative benchmark for the burden of injury in Canada and underscore the critical importance of preventive interventions. This data can inform the allocation of resources and the development of age-targeted public health strategies to reduce injury incidence and associated economic costs. Additionally, although this estimate is only a rough approximation, we believe it provides a basis for future large-scale, population-based prospective cohort studies to more accurately quantify the lifetime risk of all-cause injury-related hospitalization among Canadians.

## Conclusion

Based on this analysis, we estimate that approximately 1 in every 2.4 Canadians will be hospitalized for injury at least once in their lifetime. This estimation can be used to augment the speaking points related to injury and the urgency for substantive and coordinated prevention efforts. While injury continues to be a leading cause of death for Canadians under the age of 45 years, the cumulative risk of hospitalization across the lifespan is deeply concerning. For healthcare providers and policymakers, this data helps contextualize risk and inform the development of interventions aimed at reducing both injury incidence and associated healthcare costs. It also provides the general population with a clear reference to understand their lifetime risk of injury-related hospitalization, supporting greater awareness and engagement in prevention efforts.

## Acknowledgments

The authors acknowledge the assistance and support from staff of Sunnybrook Health Centre and department of Centre for Injure Prevention.

## Contributors

QM was involved in data analysis, interpretation of results and drafting the manuscript. BET and SF conceptualised and oversaw the project and contributed to writing and revising the manuscript. BET serves as the guarantor for the manuscript.

## Patient consent for publication

Patients and/or the public were not involved in the design, or conduct, or reporting, or dissemination plans of this research.

## Ethics approval

The study used ONLY openly available human data from public database. https://www150.statcan.gc.ca/t1/tbl1/en/tv.action?pid=9810002701 https://www.cihi.ca/en/injury-and-trauma-emergency-department-and-hospitalization-statistics

## Data availability statement

All data produced are available online at: https://www150.statcan.gc.ca/t1/tbl1/en/tv.action?pid=9810002701 https://www.cihi.ca/en/injury-and-trauma-emergency-department-and-hospitalization-statistics

